# Macrophage spatial polarity to T cells predicts prognosis in young women with luminal breast cancer

**DOI:** 10.64898/2026.05.17.26352909

**Authors:** Artur Mezheyeuski, Garazi Serna, Alfonso Martín-Bernabé, Neda Hekmati, Ioannis Zerdes, Anna Dénes, Hanna Fredholm, Siarhei Mauchanski, Xavier Guardia, Lidia Alonso, Lynn De Mey, Tony Lahoutte, Marleen Keyaerts, Joakim Lindblad, Nataša Sladoje, Fredrik Wärnberg, Malin Sund, Gunilla Rask, Charlotta Wadsten, Fredrik Pontén, Patrick Micke, Irma Fredriksson, Paolo Nuciforo

## Abstract

**Purpose:** The prognostic role of tumor-infiltrating lymphocytes in luminal breast cancer remains uncertain, partly because density-based metrics do not capture spatial interactions between immune cell subsets. We developed a density-independent spatial metric quantifying macrophage-T cell proximity and assessed its prognostic value.

**Experimental Design:** Using multiplex immunohistochemistry across three breast cancer cohorts (exploratory, n = 17; discovery, n = 687; validation, n = 305), we measured nearest-neighbor distances from T cells to M1-like and M2-like macrophages, benchmarked against a randomly subsampled total macrophage pool. We defined the Macrophage Spatial Polarity Index (MSPI) as the difference between M2-to-T cell and M1-to-T cell affinity scores, where higher values reflect an M2-dominated spatial phenotype. Cox regression was used to assess associations with distant disease-free survival (discovery) and overall survival (validation).

**Results:** M2-like macrophages preferentially localized near T cells, independent of cell density. Higher MSPI was associated with shorter survival in luminal cancers (discovery: HR = 1.45, p < 0.001), with the strongest effect in young women with early-stage disease (HR = 2.16, p < 0.0001). MSPI remained independently prognostic after adjustment for stage, systemic treatment, and diagnosis period (HR = 2.31, 95% CI 1.73-3.09, p < 0.0001) and was non-significant in HER2-positive and triple-negative subtypes. Validation in an independent ER-positive cohort confirmed the finding (HR = 1.30, p = 0.004). Pooled analysis yielded HR = 2.13 (95% CI 1.68-2.70, p = 3.45 x 10⁻¹⁰).

**Conclusions:** MSPI is a robust prognostic biomarker in luminal breast cancer, particularly in young women with early-stage disease, warranting further validation for risk stratification and therapeutic guidance.

## Introduction

The tumor microenvironment (TME) is a dynamic ecosystem in which cancer cells, stromal elements, and immune cells co-evolve, collectively shaping tumor progression and response to therapy. Tumor-infiltrating lymphocytes (TILs) are among the most studied immune components of the TME, and their clinical significance has been firmly established for specific breast cancer subtypes. In triple-negative breast cancer, a pooled analysis of > 2,000 patients demonstrated a linear dose-response relationship between stromal TIL density and improved survival after adjuvant chemotherapy (1). In HER2-positive disease, higher TIL levels predict trastuzumab benefits and a longer event-free survival (2).

However, the prognostic role of TILs in luminal hormone receptor-positive (HR-positive) breast cancer remains controversial. A pooled analysis of 3,771 patients treated with neoadjuvant therapy revealed that high TIL levels were associated with improved outcomes in HER2-positive and triple-negative disease but with worse prognosis in luminal-HER2-negative tumors (3). Emerging single-cell and spatial transcriptomic data have revealed that the immune landscape of HR-positive tumors is qualitatively distinct, with immunosuppressive macrophage subsets colocalized with T cells in tumor-associated niches (4). These findings suggest that simple immune cell counts are insufficient to capture the functional complexity of the immune infiltrate in luminal breast cancer, in which the spatial organization and polarization of immune cells may be more informative than their abundance.

These observations have gained clinical urgency with a recent demonstration that immune checkpoint inhibitors can improve the pathological response in ER-positive breast cancer. The phase 3 CheckMate 7FL trial showed that adding nivolumab to neoadjuvant chemotherapy significantly increased pathological complete response (pCR) rates in high-risk ER-positive/HER2-negative breast cancer (24.5% vs. 13.8%, p = 0.002), with the greatest benefit observed in tumors with high PD-L1 expression (pCR 44.3% vs. 20.2%) (5). Similarly, the KEYNOTE-756 trial demonstrated that pembrolizumab plus chemotherapy improved pCR compared to chemotherapy alone (24.3% vs. 15.6%, p < 0.001), with enhanced benefits in PD-L1-positive and TIL-rich tumors (6). These trials established that a subset of luminal breast cancers harbors an immune microenvironment amenable to immunotherapy, but current biomarkers, including PD-L1 expression and TIL density, provide only a partial picture of immune engagement (7). Understanding the spatial organization of immune cells, particularly the functional interactions between macrophages and T cells that regulate antitumor immunity, could help identify which luminal tumors are likely to benefit from immune-based treatment strategies (8,9).

Clinical management of luminal breast cancer in young women represents a challenge. Updated international consensus guidelines recognize young age as an independent adverse prognostic factor and recommend intensified endocrine strategies, including ovarian function suppression combined with aromatase inhibition, yet acknowledge the absence of validated biomarkers to guide treatment intensity within this population (10). Long-term follow-up of randomized trials has confirmed that premenopausal women with HR-positive breast cancer benefit from ovarian function suppression plus aromatase inhibition, with disease-free survival gains most pronounced in women who also received adjuvant chemotherapy (11). Treatment decisions, particularly regarding chemotherapy escalation, duration of endocrine therapy, addition of CDK4/6 inhibitors, and now potential immunotherapy, depend on accurate prognostic stratification. The identification of microenvironment-based markers that capture immune-tumor interactions could help refine risk assessment and guide personalized treatment decisions in this population (12,13).

Tumor-associated macrophages (TAMs) are among the most abundant immune cells in the TME of breast cancer and exhibit remarkable functional plasticity. The classical M1/M2 polarization framework provides a conceptual foundation: M1-like macrophages are generally anti-tumorigenic through pro-inflammatory cytokine production and antigen presentation, whereas M2-like macrophages promote tumor growth through immunosuppression, angiogenesis, and tissue remodeling (14). Recent work has demonstrated that TAMs in breast cancer actively restrict CD8 + T cell function through collagen deposition and metabolic reprogramming of the peritumoral stroma, establishing physical and biochemical barriers to antitumor immunity (15). Previously, we showed the importance of TIL/macrophage balance across cancers (16). However, density-based metrics alone cannot distinguish macrophages that are passively present in the TME from those that actively interact with T cells, a distinction that may be critical for understanding immune suppression. The macrophage-T cell axis is of particular interest because macrophages directly regulate T cell effector function through both contact-dependent mechanisms (PD-L1/PD-1, CD80/CTLA-4) and paracrine signaling (IL-10, TGF-β), making the spatial relationship between these cell types a potential readout of local immunosuppression that cannot be captured by density measurements alone.

Advances in multiplex immunohistochemistry (mIHC) and spatial imaging technologies have enabled analysis of the spatial organization of the TME, revealing that the topology of immune cell interactions carries prognostic and predictive information beyond what cell densities alone can provide (17,18). Imaging mass cytometry of 693 breast tumors identified recurrent TME structures in which regulatory and dysfunctional T cells predicted poor outcomes in ER-positive disease (19). In triple-negative breast cancer, spatial immunophenotyping predicts response to anti-PD-1 therapy (20). These findings underscore the importance of spatial context, but most existing spatial metrics remain influenced by overall cell density and tissue architecture, making it difficult to determine whether observed spatial patterns reflect genuine biological interactions or are simply a consequence of high cell abundance. We have previously implemented spatial metrics derived from immune cell distances from the tumor/stroma interface (21–24). However, most spatial systems are affected by the cell abundance and tissue morphology.

Here, we introduce a cell-density-independent affinity metric that quantifies the spatial proximity of macrophage subtypes (M1 and M2) to T cells by comparing the observed nearest-neighbor distances against a randomly subsampled reference macrophage pool. We integrated the opposing M1 and M2 affinity signals into a composite Macrophage Spatial Polarity Index (MSPI) and established a categorical framework (qMSPI) that classified tumors into four spatial immune phenotypes. Using three independent breast cancer cohorts, an exploratory cohort for metric development (n = 17), a population-based discovery cohort for prognostic analysis (n = 687), and an independent validation cohort (n = 305; ER-positive n = 247), we demonstrated that macrophage-to-T cell spatial polarity is a robust and reproducible prognostic biomarker with particular relevance for young women with early stage luminal breast cancer.

## Materials and methods

### Study cohorts

Three breast cancer tissue collections were used for this study. The **Exploratory cohort** comprised 17 formalin-fixed, paraffin-embedded (FFPE) whole-tissue sections from surgically resected breast carcinoma specimens obtained from 10 patients aged ≥ 18 years. These samples were collected during the Phase IIa segment of the ⁶⁸Ga-NOTA-Anti-MMR-VHH2 clinical trial (25), a study on a PET/CT imaging radiotracer that targets tumor-associated macrophages (TAMs). The cohort predominantly included luminal (HR+) breast carcinomas, one HER2-positive tumor, and one triple-negative breast carcinoma (TNBC). HER2 positivity was defined as IHC 3+ or ISH amplification according to the local diagnostic assessment (Supplementary Table S1).

The **Discovery cohort** included patients selected from a Swedish population-based registry cohort of women diagnosed 1992-2005 with primary invasive breast cancer, including all women aged <35 years (n = 471), and a random sample of women aged 35-39 (n = 200), 40-49 (n = 200), and 50-69 (n = 300) (26,27). Information on patient and tumor characteristics, treatment, and follow-up was collected from the medical records. Molecular subtypes were defined as IHC-based surrogates using ER, PR, HER2, and Ki67 expression and classified into Luminal A, Luminal B, HER2-positive (HER2-enriched and Luminal-HER2), and triple-negative (TNBC) categories according to established criteria. Tissue microarrays (TMA) were constructed from FFPE blocks, and mIHC was performed. Data from 687 stage I-IV patients aged 20-69 and comprising all major subtypes, were available for spatial analysis after mIHC (Supplementary Table S2).

The **Validation cohort** comprised a population-based breast cancer tissue microarray collection from Uppsala University Hospital and Västerås Central Hospital, Sweden. The cohort included women who were diagnosed with primary breast cancer between 1985 and 2004 (28). Tissue microarrays were constructed from FFPE surgically resected specimens. In total, 305 patients had evaluable mIHC data for spatial analysis and were classified according to estrogen receptor (ER) status (≥10% positivity) as ER-positive (n = 247) or ER-negative (n = 58). The ER-positive subset was the primary validation population. All ER-positive patients had tumors ≤15 mm (pT1) with 0–3 axillary lymph node metastases. Overall survival (OS), defined as the time from diagnosis to death from any cause, was the primary endpoint (Supplementary Table S3).

### Immuno-staining and image analysis

For the **Exploratory cohort**, 3 micrometre (μm) sections were cut from FFPE tumor samples. Next-Generation Immunohistochemistry (NGI) was employed to enable sequential chromogenic staining of multiple biomarkers in the same tissue section while preserving both antigenicity and tissue morphology (24,29). The following markers were applied: CD3 (clone 2GV6, Ventana Medical Systems, Tucson, AZ, USA), CD163 (clone MRQ-26, Cell Marque, Rocklin, CA, USA), and CD68 (clone KP-1, Ventana Medical Systems). Each staining round involved chromogenic detection followed by high-resolution digital scanning using a NanoZoomer 2.0-HT slide scanner (Hamamatsu Photonics, Hamamatsu City, Japan). Digital images from each round were aligned using the Visiopharm Image Analysis software (Visiopharm A/S, Hørsholm, Denmark). Tumor regions were manually annotated by a board-certified pathologist based on histopathological criteria. Any detectable staining was considered positive for each marker. Composite images for illustration purposes were generated using ImageJ software (National Institutes of Health, Bethesda, MD, USA).

For the **Discovery cohort**, 4 μm FFPE TMA sections were deparaffinized, rehydrated, and stained with antibodies against CD4 (clone 4B12, Agilent Technologies, Santa Clara, CA, USA), CD8 (clone C8/144B, Thermo Fisher Scientific, Waltham, MA, USA), CD68 (clone PG-M1, Agilent Technologies), CD163 (clone 10D6, Novocastra/Leica Biosystems, Newcastle, UK), and visualized using Opal 520, 570, 620, and 650 reagent packs (Akoya Biosciences, Marlborough, MA, USA). Slides were scanned using a PhenoImager HT system (Akoya Biosciences, Marlborough, MA, USA) at a resolution of 0.5 µm per pixel. InForm software (Akoya Biosciences, Marlborough, MA, USA) was used for image processing and quality control, and cell segmentation was performed based on nuclear DAPI staining. Individual marker expression was measured and extracted. Cell classification was performed in the R environment based on marker combination (co-expression) patterns as described previously (30).

For the **Validation cohort**, 4 µm FFPE TMA sections were deparaffinized, rehydrated, and stained with antibodies against CD3 (M725429-2), CD68 (M0876) ( Dako; Agilent Technologies, Santa Clara, CA, United States), and CD163 (HPA046404, Atlas Antibodies, Bromma Sweden), followed by visualization with Opal 520, 570, and 650 reagent packs, respectively. Slide digitalization and cell segmentation were performed as previously described.

Cell abundance in the discovery and validation cohorts was quantified as cell density, defined as the number of positively stained cells per analyzed tissue area (cells/mm²). The following cell classes were defined: M1-like macrophages (CD68⁺CD163⁻), M2-like macrophages (CD68⁺CD163⁺), Mx myeloid cells (CD68⁻CD163⁺), and T cells (as CD3⁺ in the Exploratory and Validation cohorts and CD4⁺ and CD8⁺ in the Discovery cohort).

### Spatial proximity metrics

To assess potential spatial interactions, cell immediate proximity (contact) analysis was performed. T cells (CD3⁺) were considered in direct contact with macrophages when at least 20% of their perimeter touched any macrophage subtype, as measured by image analysis.

The following algorithm was developed to compute the **spatial affinity metric**. For each sample, the nearest-neighbor distance (NND) between T cells and M1 macrophages (or M2 macrophages, depending on the analysis) was identified by analyzing all available cells within a radius of 50 µm. For the reference group, the total macrophage (MF) pool, including M1+M2+Mx, was used. As the entire MF pool is more numerous than the M1 or M2 subsets, random subsampling was performed to match the number of M1 (or M2) cells, ensuring a balanced comparison. Random sampling was repeated five times, and the average nearest distance value was used for the analysis.

The affinity metric ‘M1-to-T cell’ (or ‘M2-to-T cell’) is thus a ratio between the NND between M1 (or M2) and T cells, and the averaged NND between the MF pool and T cells. For survival analyses, cell-level affinities were averaged for each patient. This yielded two per-patient scores: M1-to-T cell affinity score and M2-to-T cell affinity score.

To integrate the opposing prognostic signals of M1 and M2 affinities into a single composite, we defined the **Macrophage Spatial Polarity Index (MSPI)** as the difference between the M2-to-T cell and M1-to-T cell affinity scores, such that higher values indicate an M2-dominated spatial phenotype. Continuous Cox regression was used to estimate hazard ratios (HR) per standard deviation (SD) increase in MSPI. Multivariable models were adjusted for T-cell density (log-transformed) and macrophage density (log-transformed). In addition, tumors were classified into four spatial immune phenotypes using outcome-independent median splits of the M1-to-T and M2-to-T cell affinity scores: Q1 (M1-close/M2-far, activation dominant), Q2 (both close and mixed), Q3 (both far and disengaged), and Q4 (M2-close/M1-far, suppression dominant). Cox regression was used to compare Q4 with Q1 and non-Q4.

### Statistical analysis

For the discovery cohort, distant disease-free survival (DDFS) was defined as the time from diagnosis until either distant recurrence or death from breast cancer, whichever occurred first, with patients being censored at the date of the last follow-up.

For the validation cohort, overall survival (OS) was defined as the time from diagnosis to death from any cause, with living patients censored at the last follow-up.

Because of the selective nature of the discovery cohort (relative enrichment of young patients), the young patient group was defined as those aged <40 years. The same threshold, if applied to the validation cohort, would result in too few patients, and therefore was raised to ≤50 years old.

To assess the independent prognostic contributions of M1 and M2 spatial affinity, an additive Cox proportional hazards model was fitted with M1-to-T cell and M2-to-T cell affinity scores simultaneously included as continuous covariates (untransformed, per 1-SD increase). A likelihood ratio test (LRT) was used to compare the multiplicative interaction model (M1 + M2 + M1×M2) with the additive model (M1 + M2) to evaluate whether the two signals combine additively.

Comparisons of continuous variables across three or more groups were performed using the Kruskal-Wallis test, with Dunn’s post-hoc test for pairwise comparisons adjusted for multiple testing using the Benjamini-Hochberg method. For two-group comparisons, the Wilcoxon rank-sum test was used. The chi-square test was used to evaluate the associations between the variables. Kaplan-Meier estimates were used to generate survival curves and were compared using the log-rank test. Cox proportional hazards models were used to estimate uni- and multivariable hazard ratios (HR) with corresponding 95% confidence intervals, and the Wald test was used for p-value estimation. For survival analyses, digital immune cell densities and affinity metrics were dichotomized using optimal thresholds. The Empirical Cumulative Distribution Function (ECDF) was used to visualize and compare the distributions of cell distances, with the Kolmogorov-Smirnov test to compare distributions. All statistical tests were two-sided, and a p-value <0.05 was considered statistically significant.

To quantify the overall MSPI effect across cohorts, stratified Cox regression was performed by pooling individual patient data from the discovery and validation cohorts with within-cohort standardization of the MSPI score.

All analyses were performed using RStudio (version 2024.09.0).

## Results

### M2 macrophages preferentially contact T cells in breast cancer tissue

To explore macrophage-T cell immediate proximity (direct contact), we analyzed the exploratory cohort with an IHC panel targeting T cells and M1-like and M2-like macrophages. M2-like macrophages constituted the largest proportion (48% of the identified immune cells, 26% of T cells, and 7.6% of M1-like macrophages). We observed a recurring spatial configuration in which T cells were encircled by cytoplasmic extensions of adjacent macrophages. We refer to this distinct cellular arrangement as “hugging,” where a macrophage (the “hugger”) envelops a T cell (the “hugged”) with protrusions (Fig 1b). We quantified this phenomenon by calculating the proportion of T cells in direct contact with the macrophages. Only 5.4% of the M1-like macrophages were in direct contact with T cells, whereas this fraction reached 16.1% for M2-like macrophages (Fig 1c, left panel). From the T cell perspective, only 0.8% of T cells were in direct contact with M1-like macrophages, whereas 24.6% were in direct contact with M2-like macrophages (Fig 1c, right panel).

**Figure 1.**
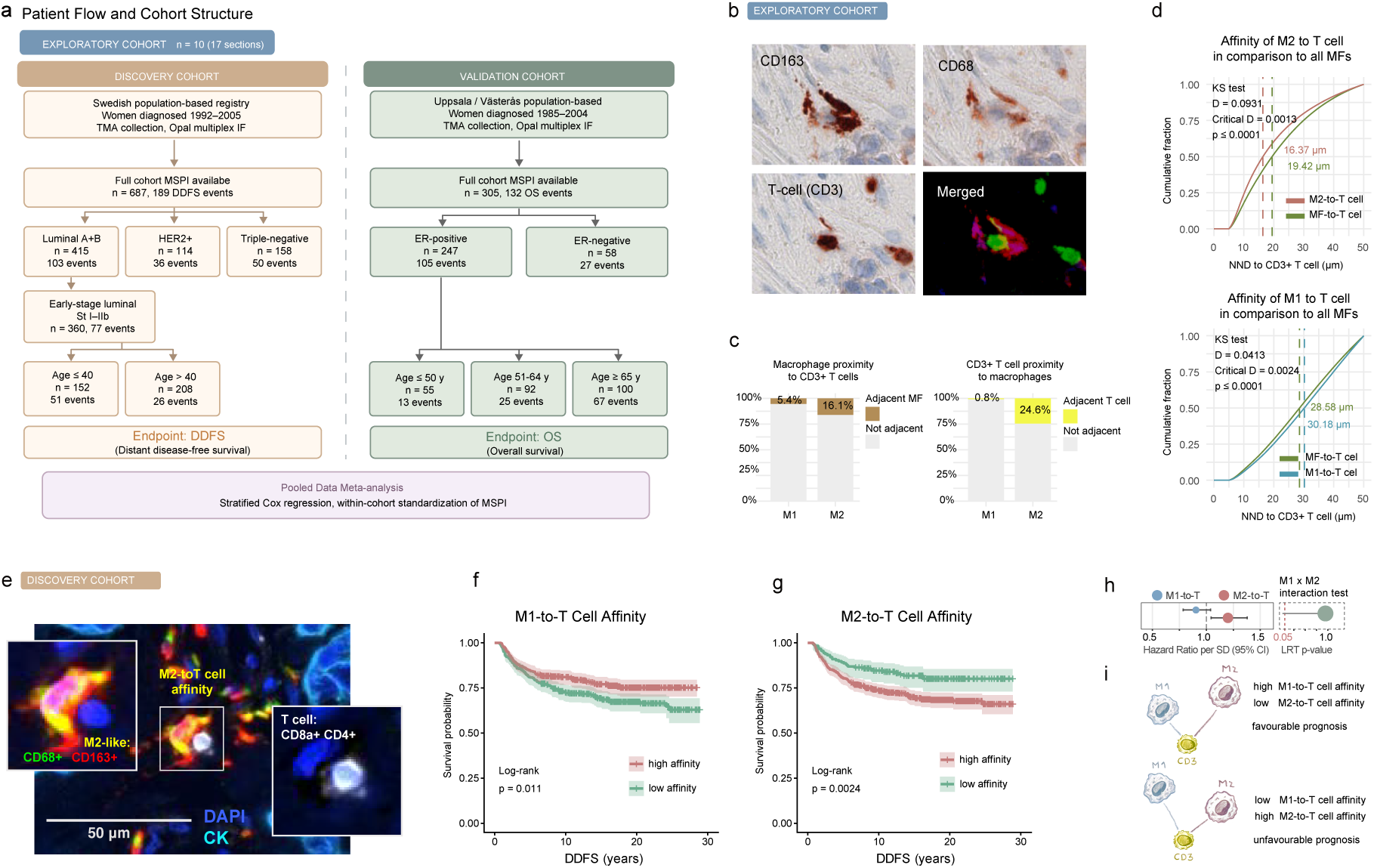
Study design, macrophage-T cell spatial relationships, and the affinity metric in breast cancer. **(a)** Patient flow and cohort structure. **(b)** Representative immunohistochemistry images from the Exploratory cohort showing CD163 (M2 macrophages), CD68 (pan-macrophage), CD3 (T cells), and the merged composite image. **(c)** Quantification of spatial adjacency in the Exploratory cohort. Left panel: proportion of macrophages within a defined radius of a CD3-positive T cell, stratified by M1 and M2 subtype. M2 macrophages showed higher adjacent fractions. Right panel: reciprocal analysis showing the proportion of T cells adjacent to macrophages. **(d)** Cumulative distribution functions of nearest-neighbor distances (NND) from macrophage subtypes to CD3-positive T cells in the Exploratory cohort. Upper panel: M2-to-T cell distances (median 16.37 μm) compared with the randomly subsampled total macrophage pool (median 19.42 μm). Two-sample Kolmogorov-Smirnov (KS) test used for statistical comparison. Lower panel: M1-to-T cell distances compared with the macrophage pool. The leftward shift of the M2 curve relative to the pool confirms that M2 macrophages are positioned closer to T cells than expected by chance, whereas M1 macrophages are positioned farther. **(e)** Representative multiplex IHC image from the Discovery cohort, showing T cells, M2 macrophages and neighboring to direct contact events. **(f-g)** Kaplan-Meier survival curves for M1-to-T cell and M2-to-T cell affinity in the Discovery cohort. Patients dichotomized at the optimal cutpoint determined by maximally selected rank statistics (with 200-iteration bootstrap validation). **(h)** Forest plot showing hazard ratios per standard deviation (HR per 1-SD increase) for M1-to-T cell affinity (blue) and M2-to-T cell affinity (terracotta) from an additive Cox proportional hazards model (as independent covariates) in the discovery cohort. The horizontal bars with vertical caps indicate 95% confidence intervals. Right panel: Likelihood ratio test (LRT) p-value comparing the multiplicative interaction model (M1 + M2 + M1 x M2) against the additive model (M1 + M2). **(i)** Schematic representation of the affinity metric concept. The affinity score compares the nearest-neighbor distance from each T cell to a macrophage subtype (M1 or M2) versus the distance to a randomly subsampled pool of all macrophages. A score below 0.5 indicates that the macrophage subtype is closer to T cells than expected by chance (high affinity). High M1-to-T cell affinity combined with low M2-to-T cell affinity defines a favorable spatial immune phenotype. The inverse pattern (low M1 affinity, high M2 affinity) defines an unfavorable phenotype. **Abbreviations:** DDFS, distant disease-free survival; ER, estrogen receptor; HR, hazard ratio; IHC, immunohistochemistry; KS, Kolmogorov-Smirnov; MSPI, Macrophage Spatial Polarity Index; N0, node-negative; NND, nearest-neighbor distance; OS, overall survival.

Next, we compared the abundance of cells in direct contact with the density of individual cell types at the per-sample level. We observed a strong correlation between contact scores and the densities of T cells and macrophages, indicating that the frequency of contact was influenced by overall cell abundance (Supplementary Figure S1, a-d).

Together, these findings show that close spatial proximity to T cells is a distinctive feature of M2-like macrophages but not M1-like macrophages in breast cancer. Although this spatial preference may reflect the underlying biological interactions between these immune cell types, it is affected by the overall cell density.

### A density-independent metric captures macrophage-T cell spatial affinity

Next, we developed a method to quantify pairwise cell-cell spatial relationships that mitigate the effects of cell density while also considering the spatial characteristics of the tissue sample. This approach isolates the intrinsic tendency for spatial proximity between two cell types by comparing to a broader ‘reference’ cell category. Thus, the affinity of M2-to-T cells can be benchmarked against that of the total macrophage pool (M1 + M2 + Mx) and MF-to-T cells.

We applied this analysis by examining the affinity of M1 and M2 cells towards T cells at distances of up to 50 µm. These results confirmed our earlier observations and demonstrated a clear positive M2-to-T cell affinity (Fig 1d, upper panel). This behavior was evident throughout the analyzed distance range, suggesting potential short-range paracrine attraction. In contrast, M1 cells showed a negative affinity towards T cells (Fig 1d, lower panel) when compared to the total macrophage pool. Importantly, the affinity metric demonstrated no statistically significant association with cell density when summarized at the sample level (Supplementary Figure S1, e-h).

In summary, we demonstrated that M2, but not M1, macrophages in breast cancer tissues displayed an increased affinity towards T cells that was independent of the total cell density.

### M2-to-T cell affinity is associated with adverse prognosis in the discovery cohort

We hypothesized that M1-to-T cell and M2-to-T cell affinity could indicate functional interactions and thus have prognostic implications. To test this hypothesis, we applied an affinity analysis to a larger discovery cohort (Fig 1a). The representative staining results are shown in Fig 1e.

Next, we investigated the prognostic value of M2-to-T cell and M1-to-T cell affinity. We transformed the affinity metrics into case-specific scores and found no strong associations between the scores and cell densities (Supplementary Figure S2). The affinity scores were binarized with the optimal cutoff for the log-rank test to allow for Kaplan-Meier visualization. In the full discovery cohort with available M1-to-T cell affinity scores, higher M1-to-T cell affinity was significantly associated with longer DDFS, indicating that close spatial proximity of M1 macrophages to T cells is protective (Fig 1f). The M2-to-T cell affinity score demonstrated an opposite prognostic association with shorter DDFS (Fig 1g), indicating that the close spatial proximity of M2 macrophages to T cells is associated with adverse outcomes.

To formally validate the independent prognostic impact of both affinity scores, we fitted Cox models including M1-to-T cell and M2-to-T cell affinity scores as independent additive terms and tested for their multiplicative interaction using LRT. We used continuous, non-transformed affinity scores showing hazard ratios per standard deviation (HR per 1-SD increase) (Fig 1h). In the additive model, M2 affinity was the dominant prognostic factor, whereas M1 affinity demonstrated a directionally consistent but non-significant contribution. The LRT p-value exceeded 0.05 (p = 0.95), confirming that M1 and M2 affinity contribute independently without synergistic amplification.

Overall, the contrasting prognostic directions of the M1-to-T cell and M2-to-T cell affinity scores, protective and adverse, respectively, suggest that M1 and M2 macrophages exert independent and opposing prognostic effects through their spatial interactions with T cells (Fig 1i).

### MSPI as a prognostic composite

Given the opposite and independent prognostic effects of the M1 and M2 spatial affinity signals, we integrated them into a single composite metric. We defined the Macrophage Spatial Polarity Index (MSPI) as the difference between the M2-to-T cell and M1-to-T cell affinity scores, such that higher values indicated an M2-dominated spatial phenotype, with M2 macrophages positioned closer to T cells and M1 macrophages relatively more distant (Fig 2a, upper panel). To enhance biological interpretability, we also established a categorical framework and classified tumors into four spatial immune phenotypes, qMSPI, using outcome-independent median splits of the M1 and M2 affinity scores: Q1, M1-close/M2-far, representing an activation-dominant state; Q2, both close, representing mixed spatial signaling; Q3, both far, representing relative immune disengagement; Q4, M2-close/M1-far, representing a suppression-dominant state (Fig 2a, lower panel). Interestingly, while the individual cell densities differed across molecular subgroups within the discovery cohort (Luminal, HER2-positive and TNBC), the MSPI showed no statistically significant difference by pairwise Dunn’s test (Benjamini-Hochberg adjusted) and only a marginal difference by Pearson’s chi-squared test, across subtypes (Fig 2b and Supplementary Table S4).

**Figure 2.**
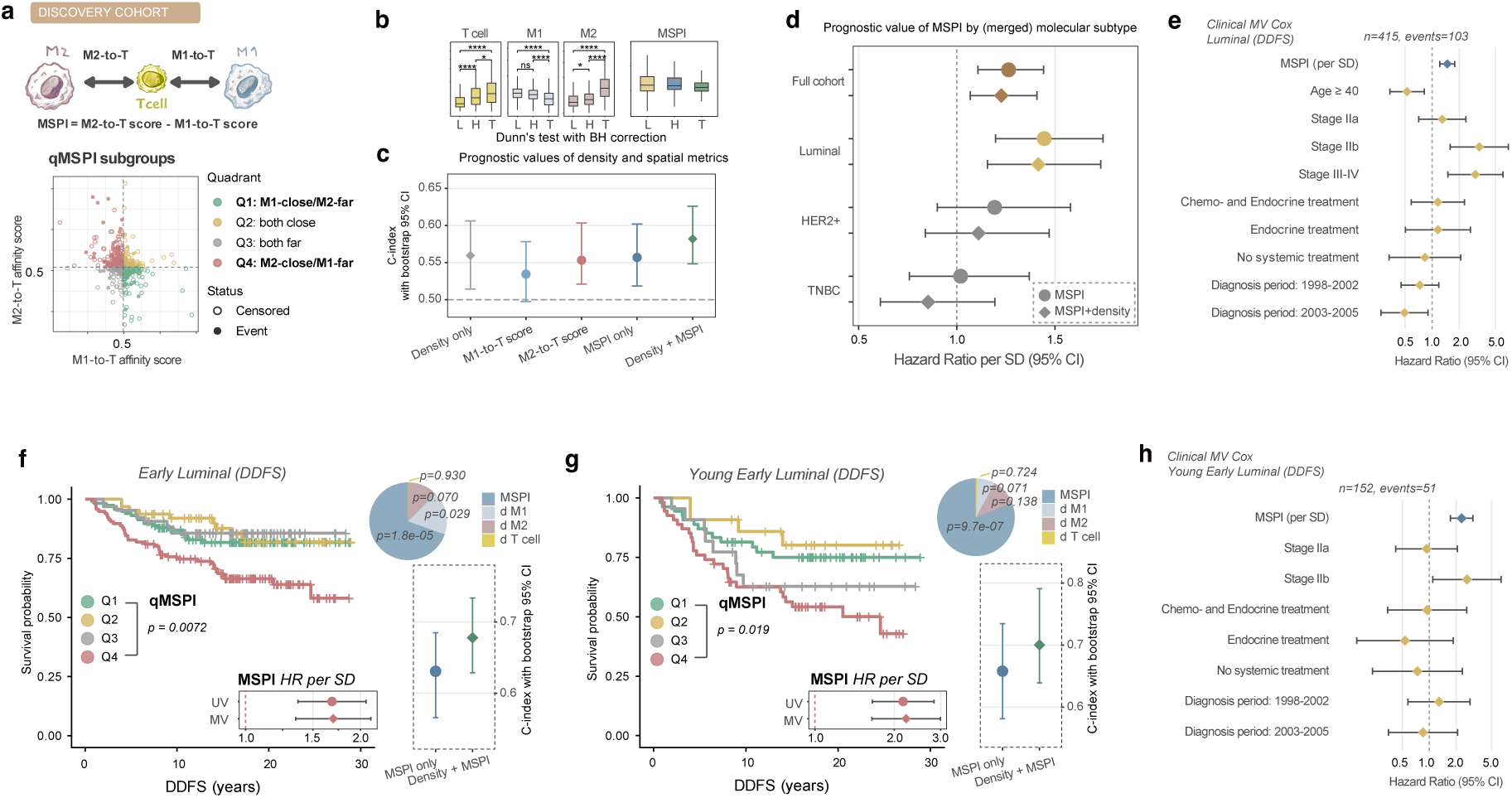
Macrophage Spatial Polarity Index (MSPI) as a prognostic composite in luminal breast cancer. **(a)** Definition and categorical framework of the MSPI. Upper panel: schematic representation of the MSPI computation. The M2-to-T cell affinity score and M1-to-T cell affinity score are combined as MSPI = M2 score - M1 score, such that higher MSPI values indicate an M2-dominated spatial immune phenotype. Lower panel: scatter plot of per-patient M1 affinity score (x-axis) versus M2 affinity score (y-axis), with tumors classified into four spatial immune phenotypes (qMSPI) using outcome-independent median splits of the M1 and M2 affinity scores. Q1: M1-close/M2-far (activation dominant); Q2: both close (mixed); Q3: both far (disengaged); Q4: M2-close/M1-far (suppression dominant). Dashed lines indicate median cutpoints. The 0.5 threshold on the x-axis corresponds to the reference point where macrophage-subtype proximity equals random expectation. **(b)** Distribution of individual cell densities and MSPI by merged molecular subtype in the Discovery cohort. Boxplots show T cell, M1 macrophage, and M2 macrophage densities (cells/mm^2^), and the composite MSPI across luminal (L), HER2-positive (H), and triple-negative (T) breast cancers (Dunn’s test with BH correction). **(c)** Comparison of prognostic capacity across density-based and spatial metrics. Harrell’s C-index with bootstrap 95% confidence intervals (200 iterations) for five Cox regression models: cell density alone (CD3 + M1 + M2 densities), M1 affinity alone, M2 affinity alone, MSPI alone, and the combined density + MSPI model. Discovery cohort, full cohort (n = 687), endpoint: DDFS. **(d)** Forest plot of MSPI hazard ratios by merged molecular subtype. Cox proportional hazards regression showing HR per 1-SD increase in MSPI for the full Discovery cohort and by molecular subtypes. Two estimates are shown per subgroup: MSPI alone (univariable, circles) and MSPI adjusted for cell densities (CD3 + M1 + M2 densities, diamonds). Horizontal bars indicate 95% confidence intervals. Endpoint: DDFS. **(e)** Clinical multivariable Cox regression for luminal breast cancer. Forest plot showing hazard ratios with 95% confidence intervals from a multivariable Cox model including MSPI (per SD, continuous), age (≥40 vs <40), TNM stage (IIa, IIb, III-IV vs I), systemic treatment (chemo and endocrine, endocrine only, no treatment vs chemo only), and diagnosis period (1998-2002, 2003-2005 vs 1992-1997). Luminal subgroup of the Discovery cohort, n = 415, 103 DDFS events. **(f)** MSPI prognostic analysis in early-stage luminal patients (stage I-IIb, n = 360, 77 DDFS events). Left panel: Kaplan-Meier curves for the four qMSPI spatial phenotypes (Q1-Q4). Below the KM curve: forest plot showing MSPI HR per SD, univariable (UV) and cell density-adjusted (MV). Right lower panel: Harrell’s C-index with bootstrap 95% CI comparing MSPI alone, and the combined cell density + MSPI model. Right upper panel: pie chart showing the relative contribution (Type II Anova chi-square proportions) of each variable in the cell density-adjusted multivariable Cox model, illustrating that MSPI accounted for the largest share of prognostic information. **(g)** MSPI prognostic analysis in young early-stage luminal patients (stage I-IIb, age <40 years, n = 152, 51 DDFS events). Layout identical to panel (f). Left panel: Kaplan-Meier curves for qMSPI phenotypes. Below the KM curve: forest plot (UV and MV). Right lower panel: C-index comparison across models. Right upper panel: chi-square contribution pie chart. **(h)** Clinical multivariable Cox regression for young early-stage luminal patients, including MSPI (per SD), stage, systemic treatment, and diagnosis period. **Abbreviations:** CI, confidence interval; DDFS, distant disease-free survival; HR, hazard ratio; KM, Kaplan-Meier; LRT, likelihood ratio test; MSPI, Macrophage Spatial Polarity Index; MV, multivariable; qMSPI, quadrant-based MSPI classification; SD, standard deviation; TNBC, triple-negative breast cancer; UV, univariable.

Next, we analyzed the prognostic capacity of several metrics derived from T cells, M1, and M2 macrophages: combined cell-density model (CD3 + M1 + M2 in a multivariable setup), M1-to-T cell and M2-to-T cell affinity scores, MSPI alone, and in combination with cell densities (as a four-variable model). The combined four-variable MSPI+cell density model outperformed other models (Fig 2c).

This demonstrates that the MSPI captures the prognostic information contained in both M1-to-T cell and M2-to-T cell affinity scores, while yielding a single interpretable score. Moreover, cell density, when combined with MSPI, improved the C-index, suggesting an independent and additive contribution of cell density and their topology to prognosis.

### MSPI in early-stage luminal and young patient subgroups

Next, we investigated whether the prognostic capacity of MSPI differs in IHC-based molecular subtypes. Interestingly, in luminal (A+B) cancers, MSPI was strongly associated with shorter DDFS, both as a single metric and adjusted for cell density (Figure 2d and Supplementary Table S5). However, in the HER2-positive cancers and in TNBC the continuous MSPI was not a prognostic. To further validate the MSPI associations in luminal cancers, we performed multivariable Cox regression analyses, including clinical covariates (age, TNM stage, systemic treatment, and diagnosis period). MSPI remained independently associated with DDFS (HR = 1.47 per SD, 95% CI 1.21-1.78, *p < 0.0001*). Among the clinical covariates, age ≥40 years was associated with favorable outcomes, while advanced stage was associated with risk (Fig 2e and Supplementary Table S6).

Given that MSPI demonstrated the strongest association with survival in luminal breast cancers, we performed a focused analysis of predefined luminal subgroups. We focused on early stage and young patients with luminal breast cancer, as treatment decisions in these groups remain particularly challenging, and robust prognostic markers are critically needed. In early-stage luminal patients (stage I-IIb), MSPI was associated with substantially worse DDFS in the univariate and multivariate models, while the C-index reached 0.631 for MSPI alone and 0.678 for the combined model with cell density (Fig 2f and Supplementary Table S7). Interestingly, the relative contribution analysis showed that MSPI accounted for the largest share of prognostic information in the density-adjusted model, followed by M1 and M2 cell densities, with negligible contribution from T cell density (Fig 2f pie-chart). The effect was further amplified in young patients with early stages (stage I-IIb, age <40), where MSPI showed the strongest survival association (density-adjusted HR = 2.22, *p < 0.0001*) and a C-index of 0.658 for MSPI alone and 0.700 for the combined model (Fig 2g and Supplementary Table S7). In the four-variable model (adjusted to cell densities), MSPI dominated the prognostic contribution even more strongly than in the early luminal group (Fig 2g pie-chart).

Finally, we performed a multivariable analysis with clinical covariates in young patients with early-stage luminal tumors (Fig 2h and Supplementary Table S8). MSPI showed the strongest independent association with DDFS (HR = 2.31 per SD, 95% CI 1.73-3.09, *p < 0.0001*), exceeding the prognostic contribution of systemic treatment and diagnosis period and being only slightly weaker than tumor stage IIb vs stage I.

In summary, MSPI showed particularly pronounced prognostic associations in young patients with early-stage luminal breast cancer, suggesting that this subgroup may benefit most from spatial immune profiling.

### Validation of MSPI in the independent cohort

To validate the prognostic significance of MSPI in luminal cancers, we applied the same analytical framework to the validation cohort (Supplementary Table S9). As the closest available approximation to the luminal AB subtype, we selected the ER-positive subset of cancers in the validation cohort and analyzed overall survival as the endpoint, as DDFS data were not available for this cohort (n = 247, 105 events). A higher MSPI was significantly associated with shorter OS in the ER-positive group (HR = 1.30 per SD, 95% CI 1.09-1.55, p = 0.004; density-adjusted HR = 1.27, p = 0.008) (Fig 3a and Supplementary Table S10). No significant association was observed in the ER-negative subset or the fully unselected cohort, confirming that the MSPI signal is specific to ER-positive/luminal disease. Interestingly, despite this dissimilarity in survival associations, MSPI values did not differ between ER-positive and ER-negative cancers, reflecting the observation in the discovery cohort (Fig 3b and compare with Fig 2b).

**Figure 3.**
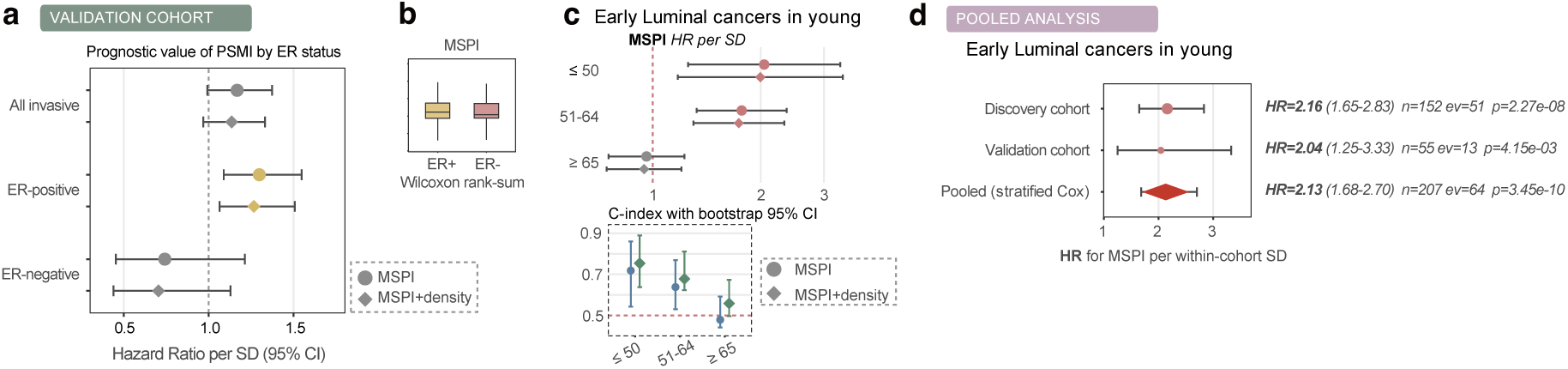
Independent validation and pooled analysis of MSPI in ER-positive breast cancer (Validation cohort, OS). **(a)** MSPI and overall survival by estrogen receptor (ER) status in the Validation cohort. Forest plot showing hazard ratios per 1-SD increase in MSPI for three subgroups: all invasive tumors (n = 305, 132 events), ER-positive (n = 247, 105 events), and ER-negative (n = 58, 27 events). Two estimates are shown per subgroup: MSPI alone (univariable, circles) and MSPI adjusted for cell densities (CD3 + M1 + M2 densities, diamonds). The dashed vertical line indicates HR = 1.0 (no effect). Endpoint: OS. **(b)** Distribution of MSPI by ER status. Boxplots comparing MSPI values between ER-positive (n = 247) and ER-negative (n = 58) tumors in the Validation cohort (Wilcoxon rank-sum). **(c)** Age-stratified MSPI prognostic performance in ER-positive patients from the Validation cohort. Upper panel: forest plot of MSPI hazard ratios per SD for three age groups: ≤50 years (n = 55, 13 events), 51-64 years (n = 92, 25 events), and ≥65 years (n = 100, 67 events). Two estimates per subgroup: univariable (circles) and density-adjusted (diamonds). Lower panel: Harrell’s C-index with bootstrap 95% confidence intervals (200 iterations) for MSPI alone (circles) and the combined cell density + MSPI model (diamonds) in each age group. Endpoint: OS. **(d)** Pooled individual patient data meta-analysis for MSPI in early luminal cancers in young patients. Forest plot showing MSPI hazard ratios per within-cohort SD from the Discovery cohort (luminal stage I-IIa, age <40), the Validation cohort (ER-positive, age ≤50), and the pooled estimate from a stratified Cox regression. The stratified Cox model accounts for between-cohort differences in baseline hazard while estimating a common MSPI effect. The diamond represents the pooled estimate with 95% confidence interval. **Abbreviations:** CI, confidence interval; DDFS, distant disease-free survival; ER, estrogen receptor; HR, hazard ratio; MSPI, Macrophage Spatial Polarity Index; MV, multivariable; N0, node-negative; OS, overall survival; SD, standard deviation; UV, univariable.

Next, we intended to validate the superior performance of MSPI in young patients with early-stage tumors. The validation cohort consisted of patients with low-stage tumors by design. To mirror the age-stratified analysis performed in the discovery cohort, we examined the MSPI in ER-positive patients stratified by age groups available in this dataset: ≤50 years (n = 55, 13 OS events), 51-64 years (n = 92, 25 events), and ≥65 years (n = 100, 67 events). In patients aged ≤50 years, MSPI showed the strongest effect (HR = 2.04 per SD, 95% CI 1.25-3.33, *p = 0.004*; density-adjusted HR = 1.99, *p = 0.011*), with good prognostic discrimination (C-index: MSPI alone = 0.72, density + MSPI = 0.75). In the 51-64 age group, the MSPI effect remained significant, but the magnitude was weaker. In patients aged ≥65 years, MSPI showed no significant association, and the C-index was near 0.50, indicating no prognostic value in this age group (Fig 3c and Supplementary Table S11).

### Pooled individual patient data meta-analysis

To formally quantify the combined evidence across cohorts, we performed a pooled meta-analysis using stratified Cox regression with within-cohort standardization of MSPI scores. We pooled the discovery of young early stage luminal (stage I-IIb, age <40 years) and validation of ER-positive young node-negative (pT1, axN0-3, age ≤50 years) patients. The MSPI effect was consistent across both cohorts: HR = 2.16 per SD in the validation cohort (hazard HR = 2.04 in the validation cohort. The pooled estimate yielded HR = 2.13 per SD (95% CI 1.68-2.70, *p = 3.45 x 10-10*; n = 207, 64 events) (Fig 3d and Supplementary Table S12).

The consistent magnitude of the MSPI effect across cohorts with different endpoints (DDFS vs. OS) and age definitions (<40 vs. ≤50 years) confirms the robustness of the MSPI as a prognostic marker in early luminal breast cancer in young patients.

## Discussion

In this study, we introduced a cell-density-independent spatial affinity metric and its composite derivative, the Macrophage Spatial Polarity Index, which captures the opposing spatial relationships of M1-like and M2-like macrophages with T cells in breast cancer tissue. Across two independent cohorts, MSPI emerged as a robust prognostic biomarker in luminal/ER-positive breast cancer, with the strongest and most clinically relevant associations observed in young women with early-stage disease. The finding was validated in an independent cohort with a different endpoint (DDFS vs OS), a different age threshold (<40 vs ≤50 years), yet the effect size remained remarkably consistent (pooled HR = 2.13 per SD, p = 3.45 × 10⁻¹⁰).

A key methodological contribution of this study is the design of a spatial metric that is inherently independent of cell density. Most existing spatial analysis approaches, including nearest-neighbor distances, co-localization indices, and Ripley’s K-function, are confounded by the fact that cells in denser tissues are closer together, regardless of any specific biological interaction (17). Our affinity metric addresses this by benchmarking the nearest-neighbor distance from each T cell to a macrophage subtype (M1 or M2) against the distance to a randomly subsampled pool of all macrophages. This internal reference controls the local cell density and tissue architecture in a single ratio, isolating the intrinsic tendency of spatial proximity. Repeated random subsampling stabilizes the estimate without excessive computational burden, and the entire analysis runs on standard nearest-neighbor algorithms (in R or Python), making it applicable to routine mIHC data from tissue microarrays without specialized infrastructure.

The choice to focus on macrophage-to-T cell rather than tumor-to-T cell spatial relationships was motivated by biological and methodological considerations. While tumor-T cell distance reflects the degree of immune infiltration into the tumor parenchyma, this information is largely captured by T cell density, which is already well studied and routinely quantifiable. In contrast, macrophage-T cell proximity captures a distinct biological process: the regulatory interaction between antigen-presenting/immunomodulatory cells and effector lymphocytes. Macrophages are the principal regulators of T-cell function in the TME and can activate (M1-like) and suppressing (M2-like) T-cell responses through direct cell contact. Quantifying this specific interaction therefore provides information about the quality of the immune response, which is independent of and complementary to the quantity of immune infiltration.”

An important observation is that MSPI and cell density provide complementary prognostic information. The combined density + MSPI model consistently yielded higher C-indices than either metric alone across all the subgroups. The chi-square contribution analysis further showed that MSPI accounted for the largest share of prognostic information in the density-adjusted model, with M1 and M2 macrophage densities contributing additional predictive power, and T cell density contributing minimally. This finding aligns with the growing recognition that spatial organization captures biology distinct from cell abundance (17,19) and suggests that integrating spatial and density-based metrics may offer a more complete characterization of the immune microenvironment than either approach alone.

A striking feature of MSPI is its subtype specificity. The index was strongly prognostic in luminal breast cancers, but showed no association with outcome in HER2-positive or triple-negative disease, despite comparable MSPI distributions across subtypes. This dissociation between the spatial phenotype and its prognostic impact suggests that the biological consequences of macrophage-to-T cell proximity differ according to tumor context. In luminal cancers, where the immune microenvironment is typically less inflamed and TIL density carries ambiguous prognostic significance, the spatial arrangement of macrophages relative to T cells may serve as a more sensitive readout of immune suppression than cell counts alone. In contrast, in triple-negative and HER2-positive tumors, higher overall immune infiltration and different immune editing mechanisms may dilute or override the specific macrophage-to-T-cell spatial signal. This subtype-specific prognostic pattern is particularly relevant considering recent immunotherapy trials on ER-positive breast cancer. In both CheckMate 7FL and KEYNOTE-756, the greatest benefit from checkpoint inhibitor addition was observed in tumors with high PD-L1 expression and TIL density features, which may reflect the same underlying biology captured by MSPI. Tumors with high M2-to-T cell spatial affinity (high MSPI) may represent a subset of luminal cancers, where active immunosuppression occurs at the macrophage-T cell interface, potentially amenable to checkpoint inhibitor-based strategies.

The amplified prognostic effect of MSPI in young patients is clinically noteworthy. International consensus guidelines identify young age as a key adverse prognostic factor and recommend treatment intensification for young women with luminal breast cancer, including ovarian function suppression, aromatase inhibition, and consideration of CDK4/6 inhibitors (10,11). However, these recommendations are broadly applied, and the absence of microenvironment-based biomarkers limits individualized risk stratification. In our discovery cohort, the MSPI in the young early-stage luminal subgroup (HR = 2.31 per SD) exceeded the prognostic contribution of systemic treatment, diagnosis period, and most clinical covariates. This was independently confirmed in the validation cohort, in which young ER-positive patients (≤50 years old) showed an MSPI-based C-index of 0.72. These data suggest that MSPI could complement existing clinical and genomic tools to identify young luminal patients at elevated risk who might benefit from intensified surveillance or treatment escalation while sparing low-risk patients from unnecessary overtreatment.

The biological basis of M2-to-T cell spatial affinity likely involves contact-dependent immunosuppressive signaling. Recent studies have shown that tumor-associated macrophages drive T cell exhaustion through sustained type I interferon signaling and direct cell-cell interactions in the tumor microenvironment (31). The functional diversity of macrophage states in cancer, now mapped at single-cell resolution, extends well beyond the M1/M2 dichotomy with distinct transcriptional programs linked to immunosuppression, angiogenesis, and tissue remodeling (32). The “hugging” phenomenon we observed in the exploratory cohort, where macrophage cytoplasmic extensions encircle T cells, is consistent with sustained cell-cell signaling that may promote T cell exhaustion or anergy. This spatial pattern may reflect a local immunosuppressive niche in which M2 macrophages actively restrain T cell cytotoxicity, a mechanism distinct from and complementary to global suppression captured by M2 cell density alone. The opposing M1 affinity signal, where closer M1-to-T cell proximity was protective, further supports a model in which the macrophage polarization state determines whether spatial proximity to T cells results in immune activation or suppression.

These findings have implications for developing macrophage-targeted therapeutic strategies. The macrophage-directed drug development landscape has expanded considerably, encompassing not only CSF1R inhibitors and macrophage reprogramming agents, but also CD47-SIRPα checkpoint inhibitors and engineered chimeric antigen receptor macrophages (CAR-M) (33). Notably, second-generation CAR-M approaches have demonstrated the ability to remodel the immunosuppressive tumor microenvironment and enhance T cell-mediated antitumor responses in preclinical models (34). Our data suggest that MSPI could serve as a patient-selection biomarker for such approaches, identifying tumors with active M2-mediated T cell suppression that might benefit from macrophage reprogramming or depletion. The qMSPI framework further enables clinical translation by categorizing patients into biologically interpretable groups: Q4 (suppression-dominant) patients represent the highest-priority candidates for interventions disrupting M2-T cell crosstalk, whereas Q1 (activation-dominant) patients may benefit from effective anti-tumor immune surveillance.

This study had several limitations. First, the study relied on mIHC, which captures only a limited number of markers and does not provide functional or transcriptomic information. The M1/M2 dichotomy based on CD68/CD163 co-expression simplifies the macrophage polarization spectrum. Second, the discovery cohort included patients diagnosed between 1992 and 2005, and treatment patterns have evolved since then, potentially affecting the generalizability of the survival associations. Third, the validation cohort was restricted to small tumors (pT1) and lacked treatment data, precluding multivariable clinical adjustment. Finally, it remains unclear whether the M2-to-T cell spatial affinity reflects direct cell-cell signaling or is driven by higher-order tissue organization, such as tertiary lymphoid structures or perivascular niches.

A related limitation is that our mIHC approach captures cell phenotype and position, but not the transcriptional state or signaling activity. Integrating the affinity metric with high-resolution spatial transcriptomics and multiplexed imaging platforms (35,36) could provide mechanistic insight into the molecular programs active at the M2-T cell interface. Specifically, subcellular spatial molecular imaging could determine whether T cells in close proximity to M2 macrophages express exhaustion markers (PD-1, TIM-3, and LAG-3) at higher levels than their spatially distant counterparts, and whether the “hugging” macrophages upregulate specific immunosuppressive ligands.

In conclusion, we presented a density-independent spatial metric and its composite index (MSPI) that captures macrophage-T cell spatial polarity in breast cancer. MSPI was consistently prognostic in luminal/ER-positive disease across two independent cohorts with different endpoints, age thresholds, and geographic populations, with the strongest effects in the clinically challenging subgroup of young women with early-stage tumors. The metric proved robust across different tissue formats (whole sections and tissue microarrays), staining panels, and imaging platforms, and captured biology complementary to cell density. These findings suggest that macrophage-to-T cell spatial organization is a promising prognostic tool warranting further validation and is a potential therapeutic target in luminal breast cancer.

## Data Availability

Data regarding the methodology, image analysis, curation and data processing, and raw data of cell classes and coordinates are available from the corresponding author upon request. The analysis code used to generate all the figures and tables reported in this study is publicly available at https://github.com/ArturMez/MSPI-breast-cancer.

## Acknowledgements

We express our gratitude to Yasmine De Maeyer and Odrade Gondy for their logistic work, patient inclusion, and data management; Gratienne Van Holsbeeck, Martine Van den Broeck, and Steffi De Triff for the radiotracer preparations; the Central Biobank UZ Brussel for the processing and storage of blood samples; and nurses Nadine Eersels, Carl Van Halewijn, Nele Risch, Cedric Roels, Petra Aalders, Brecht Segers, Kathleen Op Debeeck, and Tjibbe De Haan for patient care.

## Ethical approval

All procedures performed in this study involving human participants were in accordance with the ethical standards and with the 1964 Helsinki Declaration and its later amendments or comparable ethical standards. The protocol of the prospective open-label phase II study (NCT04168528) was approved by the Belgian Federal Agency for Medicines and Health Products, local ethics committee, and Federal Agency for Nuclear Control.

The study was approved by the Research Ethics Committee at Karolinska Institute, Stockholm, Sweden (approval number 2009/1174-31/1 and 2010/586-32) and the Regional Ethics Committee of Uppsala (Record No. 99 422, 2005:118, and 2005:118/2) for the discovery and validation cohorts, respectively.

## CRediT author statement

**Artur Mezheyeuski:** conceptualization, methodology, software, formal analysis, investigation, data curation, visualization, supervision, project administration, funding acquisition, writing - original draft, writing - reviewing and editing

**Garazi Serna**: methodology, software, writing - original draft, investigation, formal analysis, data curation, visualization, writing - reviewing and editing

**Alfonso Martín-Bernabé:** conceptualization, methodology, formal analysis, investigation, data curation, visualization, writing - original draft, writing - reviewing and editing

**Neda Hekmati:** formal analysis, methodology, software, data curation, writing - reviewing and editing

**Ioannis Zerdes:** conceptualization, methodology, software, formal analysis, investigation, writing - original draft, writing - reviewing and editing

**Anna Dénes:** methodology, writing - reviewing and editing

**Hanna Fredholm:** validation, investigation, funding acquisition, writing - reviewing and editing

**Siarhei Mauchanski:** data curation, writing - reviewing and editing

**Xavier Guardia:** data curation

**Lidia Alonso:** data curation

**Lynn De Mey:** validation, data curation, writing - reviewing and editing

**Tony Lahoutte:** validation, resources, funding acquisition, writing - reviewing and editing

**Marleen Keyaerts:** validation, resources, funding acquisition, writing - reviewing and editing

**Malin Sund:** resources, investigation, methodology, writing - reviewing and editing

**Gunilla Rask:** investigation, data curation, resources, writing - reviewing and editing

**Charlotta Wadsten:** resources, investigation, data curation, writing - reviewing and editing

**Joakim Lindblad:** methodology, investigation, funding acquisition, writing - reviewing and editing

**Nataša Sladoje:** methodology, investigation, funding acquisition, writing - reviewing and editing

**Fredrik Wärnberg:** resources, investigation, writing - reviewing and editing

**Fredrik Pontén:** resources, supervision, funding acquisition, writing - reviewing and editing

**Patrick Micke:** validation, resources, supervision, funding acquisition, writing - reviewing and editing

**Irma Fredriksson:** validation, resources, supervision, funding acquisition, writing - reviewing and editing

**Paolo Nuciforo:** validation, resources, supervision, writing - reviewing and editing

