## Supplementary material for "Macrophage spatial polarity to T cells predicts prognosis in young women with luminal breast cancer"

### Supplementary Materials

### Supplementary Table S1. Clinicopathological characteristics of patients in the exploratory cohort.

| Patient number | Age | Sex | HER2 status at diagnosis | Subtype | ER | PR | TNM staging at diagnosis |
|---|---|---|---|---|---|---|---|
| 1 | 46-50 | F | neg | Luminal A | pos | pos | cT1cN0mx |
| 2 | 61-65 | F | neg | Luminal B | pos | pos | cT1cN0 |
| 3 | 71-75 | F | pos | Luminal B | pos | pos | cT1N0 |
| 4 | 51-55 | F | neg | Luminal A | pos | pos | cT3N1 |
| 5 | 66-70 | F | neg | Luminal A | pos | pos | cT2N0 |
| 6 | 36-40 | F | pos | Luminal B | pos | pos | cT2N0 |
| 7 | 61-65 | M | neg | Luminal A | pos | pos | cT2N0 |
| 8 | 56-60 | F | neg | TNBC | neg | neg | cT1N0M0 |
| 9 | 66-70 | F | neg | Luminal B | pos | pos | cT2N1 |
| 10 | 51-55 | F | neg | Luminal A | pos | pos | cT1cN0 |

### Supplementary Table S2. Distribution of clinicopathological parameters in the discovery cohort (MSPI-available subset, N = 687).

| Characteristics | Cases (percentage)<br>N = 687 |
|---|---|
| **Age** | |
| <40 | 396 (57.6) |
| ≥40 | 291 (42.4) |
| **Stage** | |
| Stage I | 298 (43.4) |
| Stage IIa | 199 (29.0) |
| Stage IIb | 92 (13.4) |
| Stage III-IV | 97 (14.1) |
| Stage missing | 1 (0.1) |
| **Grade** | |
| Grade 1 | 93 (13.5) |
| Grade 2 | 252 (36.7) |
| Grade 3 | 339 (49.3) |
| Grade missing | 3 (0.4) |
| **Molecular subtype** | |
| Lum A | 198 (28.8) |
| Lum B | 217 (31.6) |
| Lum Her2 | 60 (8.7) |
| Her2 exp | 54 (7.9) |
| Triple neg | 158 (23.0) |
| **Systemic treatment** | |
| Chemotherapy | 188 (27.4) |
| Chemotherapy and Endocrine treatment | 212 (30.9) |
| Endocrine treatment | 167 (24.3) |

| Characteristics | Cases (percentage)<br>N = 687 |
|---|---|
| No systemic treatment | 119 (17.3) |
| Missing | 1 (0.1) |
| **Diagnosis period** | |
| 1992-1997 | 225 (32.8) |
| 1998-2002 | 286 (41.6) |
| 2003-2005 | 176 (25.6) |

**Supplementary Table S3. Distribution of clinicopathological parameters in the validation cohort (MSPI-available subset, all invasive, N = 305).**

| Characteristics | Cases (percentage)<br>N = 305 |
|---|---|
| **Age** | |
| Mean age, years | 60.0 |
| ≤50 | 73 (23.9) |
| 49–64 | 113 (37.0) |
| ≥65 | 119 (39.0) |
| **ER status** | |
| ER-positive | 247 (81.0) |
| ER-negative | 58 (19.0) |
| **Grade** | |
| Grade 1 | 140 (45.9) |
| Grade 2 | 136 (44.6) |
| Grade 3 | 29 (9.5) |
| **Nodal status** | |
| Node-negative (N0) | 253 (83.0) |
| Node-positive | 52 (17.0) |
| **Tumor size** | |
| Mean size, mm | 10.1 |
| ≤10 mm | 190 (62.3) |
| 11-20 mm | 115 (37.7) |
| >20 mm | 0 (0.0) |
| **Surgery** | |
| Mastectomy | 22 (7.2) |
| Breast-conserving | 281 (92.1) |

| Characteristics | Cases (percentage)<br>N = 305 |
|---|---|
| **Radiotherapy** | |
| Yes | 249 (81.6) |
| No | 56 (18.4) |
| **Chemotherapy** | |
| Yes | 33 (10.8) |
| No | 272 (89.2) |
| **Endocrine therapy** | |
| Yes | 79 (25.9) |
| No | 225 (73.8) |
| Unknown | 1 (0.3) |
| **Diagnosis period** | |
| 1985-1989 | 45 (14.8) |
| 1990-1994 | 165 (54.1) |
| 1995-1999 | 21 (6.9) |
| 2000-2004 | 74 (24.3) |

### Supplementary Figure S1. Relationship between cell density and macrophage-T cell contact fractions versus affinity scores in the exploratory cohort.

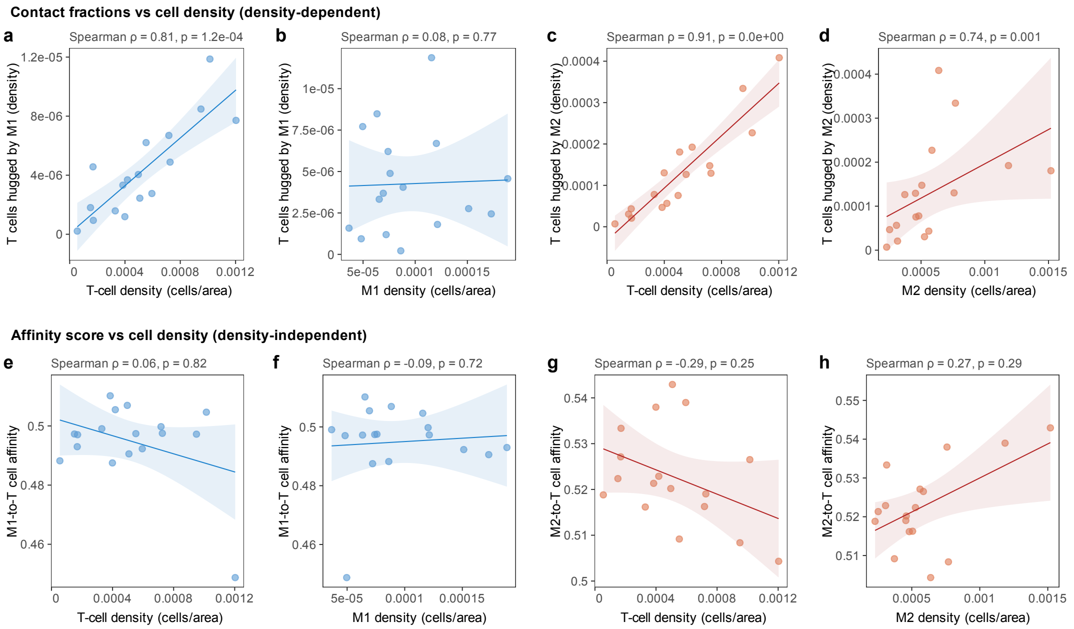


(a-d) Contact fractions versus cell density (density-dependent analysis). Scatter plots showing the relationship between cell density and the proportion of T cells in direct contact with macrophage subtypes. (a) T cells hugged by M1-like macrophages (density) as a function of T cell density (Spearman rho = 0.81, p = 1.2 x 10-4). (b) T cells hugged by M1-like macrophages (density) as a function of M1 macrophage density (Spearman rho = 0.08, p = 0.77). (c) T cells hugged by M2-like macrophages (density) as a function of T cell density (Spearman rho = 0.91, p < 0.001). (d) T cells hugged by M2-like macrophages (density) as a function of M2 macrophage density (Spearman rho = 0.74, p = 0.001). Contact fractions are strongly correlated with T cell density (panels a, c), confirming that proximity counts are confounded by cell abundance. (e-h) Affinity score versus cell density (density-independent analysis). Scatter plots demonstrating that the affinity metric is independent of cell density. (e) M1-to-T cell affinity as a function of T cell density (Spearman rho = 0.06, p = 0.82). (f) M1-to-T cell affinity as a function of M1 macrophage density (Spearman rho = -0.09, p = 0.72). (g) M2-to-T cell affinity as a function of T cell density (Spearman rho = -0.29, p = 0.25). (h) M2-to-T cell affinity as a function of M2 macrophage density (Spearman rho = 0.27, p = 0.29). In contrast to contact fractions (panels a-d), the affinity scores show no statistically significant association with cell density, confirming that the metric captures spatial proximity independent of cell abundance. Each dot represents one sample (n = 17). Lines represent linear regression fits with 95% confidence bands.

**Supplementary Figure S2. Relationship between cell density and macrophage-T cell affinity scores in the discovery cohort.**

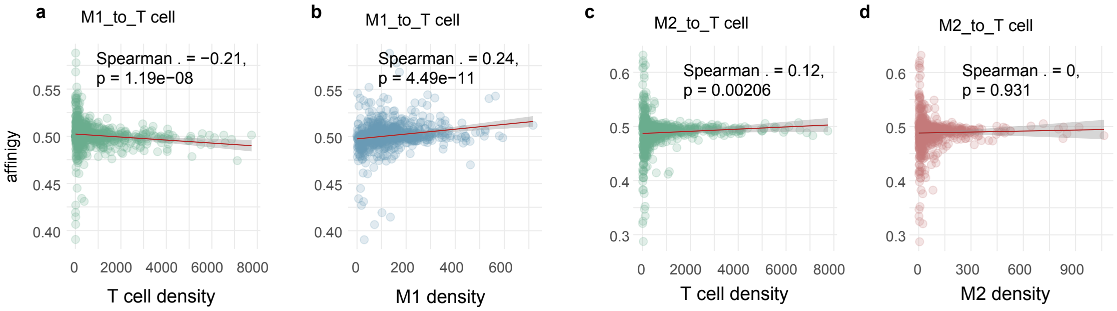


(a) M1-to-T cell affinity score as a function of T cell density (Spearman rho = -0.21, p = 1.19 x 10-8). (b) M1-to-T cell affinity score as a function of M1 macrophage density (Spearman rho = 0.24, p = 4.49 x 10-11). (c) M2-to-T cell affinity score as a function of T cell density (Spearman rho = 0.12, p = 0.002). (d) M2-to-T cell affinity score as a function of M2 macrophage density (Spearman rho = 0, p = 0.931). Although some statistically significant correlations are observed due to the large sample size (panels a-c), the correlation coefficients are weak (rho values ranging from -0.21 to 0.24), indicating that affinity scores are not strongly driven by cell density. The near-zero correlation between M2-to-T cell affinity and M2 density (panel d, rho = 0) further confirms the density-independent nature of the affinity metric. Red lines represent linear regression fits with 95% confidence bands. Discovery cohort, n = 687.

### Supplementary Table S4. Distribution of clinicopathological features by qMSPI quadrant in the discovery cohort.

| Characteristic | Q1 Best<br>N = 211[1] | Q2 Mixed<br>N = 133[1] | Q3 Interm<br>N = 132[1] | Q4 Worst<br>N = 211[1] | p-value[2] |
|---|---|---|---|---|---|
| **Age** | | | | | 0.4 |
| <40 | 130 (61.6%) | 77 (57.9%) | 70 (53.0%) | 119 (56.4%) | |
| ≥40 | 81 (38.4%) | 56 (42.1%) | 62 (47.0%) | 92 (43.6%) | |
| **TNM stage** | | | | | 0.7 |
| Stage I | 92 (43.6%) | 51 (38.3%) | 54 (40.9%) | 101 (47.9%) | |
| Stage IIa | 61 (28.9%) | 41 (30.8%) | 37 (28.0%) | 60 (28.4%) | |
| Stage IIb | 29 (13.7%) | 20 (15.0%) | 20 (15.2%) | 23 (10.9%) | |
| Stage III-IV | 29 (13.7%) | 21 (15.8%) | 20 (15.2%) | 27 (12.8%) | |
| Stage missing | 0 (0.0%) | 0 (0.0%) | 1 (0.8%) | 0 (0.0%) | |
| **Molecular subtype** | | | | | **0.037** |
| Luminal | 114 (54.0%) | 74 (55.6%) | 82 (62.1%) | 145 (68.7%) | |
| HER2+ | 42 (19.9%) | 22 (16.5%) | 18 (13.6%) | 32 (15.2%) | |
| TNBC | 55 (26.1%) | 37 (27.8%) | 32 (24.2%) | 34 (16.1%) | |
| **Systemic treatment** | | | | | **0.007** |
| No systemic treatment | 39 (18.5%) | 17 (12.9%) | 16 (12.1%) | 47 (22.3%) | |
| Endocrine treatment | 47 (22.3%) | 32 (24.2%) | 31 (23.5%) | 57 (27.0%) | |
| Chemotherapy | 72 (34.1%) | 38 (28.8%) | 41 (31.1%) | 37 (17.5%) | |
| Chemotherapy and Endocrine treatment | 53 (25.1%) | 45 (34.1%) | 44 (33.3%) | 70 (33.2%) | |
| Unknown | 0 | 1 | 0 | 0 | |
| **Diagnosis period** | | | | | 0.3 |
| 1992-1997 | 63 (29.9%) | 39 (29.3%) | 40 (30.3%) | 83 (39.3%) | |
| 1998-2002 | 88 (41.7%) | 56 (42.1%) | 60 (45.5%) | 82 (38.9%) | |
| 2003-2005 | 60 (28.4%) | 38 (28.6%) | 32 (24.2%) | 46 (21.8%) | |

[1]n (%)

[2]Pearson's Chi-squared test

**Supplementary Table S5. MSPI hazard ratios by molecular subtype in the discovery cohort.**

| Subgroup | N | Events | UV HR (95% CI) | UV p | MV HR (95% CI) | MV p |
|---|---|---|---|---|---|---|
| All-comers | 687 | 189 | 1.26 (1.11-1.44) | <0.001 | 1.23 (1.07-1.41) | 0.004 |
| Luminal | 415 | 103 | 1.45 (1.20-1.74) | <0.001 | 1.42 (1.16-1.73) | <0.001 |
| HER2+ | 114 | 36 | 1.19 (0.90-1.58) | 0.219 | 1.11 (0.84-1.47) | 0.462 |
| TNBC | 158 | 50 | 1.02 (0.76-1.37) | 0.900 | 0.85 (0.61-1.19) | 0.356 |

### Supplementary Table S6. Clinical multivariable Cox regression for MSPI in luminal breast cancer (discovery cohort).

| Varialbe | HR | CI_lo | CI_hi | p | p_fmt |
|---|---|---|---|---|---|
| MSPI (per SD) | 1.470 | 1.213 | 1.781 | 0.00008336429 | 8.34e-05 |
| Age: ≥40 | 0.526 | 0.339 | 0.815 | 0.00401454866 | 4.01e-03 |
| Stage: Stage IIa | 1.292 | 0.708 | 2.358 | 0.40310149319 | 4.03e-01 |
| Stage: Stage IIb | 3.323 | 1.579 | 6.993 | 0.00156253407 | 1.56e-03 |
| Stage: Stage III-IV | 3.007 | 1.489 | 6.074 | 0.00213857219 | 2.14e-03 |
| Systemic treatment: Chemotherapy and Endocrine treatment | 1.153 | 0.584 | 2.275 | 0.68149572780 | 6.81e-01 |
| Systemic treatment: Endocrine treatment | 1.157 | 0.504 | 2.655 | 0.73078988227 | 7.31e-01 |
| Systemic treatment: No systemic treatment | 0.831 | 0.333 | 2.074 | 0.69231727381 | 6.92e-01 |
| Diagnosis period: 1998-2002 | 0.730 | 0.451 | 1.183 | 0.20119956450 | 2.01e-01 |
| Diagnosis period: 2003-2005 | 0.493 | 0.271 | 0.897 | 0.02054589849 | 2.05e-02 |

**Supplementary Table S7. MSPI prognostic performance in early-stage luminal breast cancer (discovery cohort).**

Discovery Early Luminal | DDFS

| Model | HR | CI_lo | CI_hi | p | sig | p_fmt |
|---|---|---|---|---|---|---|
| Univariable | 1.683905 | 1.371267 | 2.067823 | 0.000000659082 | Significant | 6.59e-07 |
| Density-adjusted | 1.696765 | 1.352688 | 2.128364 | 0.000004817991 | Significant | 4.82e-06 |

Discovery Early Luminal | MSPI-focused C-index

| Model | C | CI_lo | CI_hi | AIC |
|---|---|---|---|---|
| MSPI only | 0.6309 | 0.5659 | 0.6848 | 851.2 |
| Density + MSPI | 0.6779 | 0.6289 | 0.7336 | 847.5 |

**Supplementary Table S8. MSPI prognostic performance and clinical multivariable analysis in young early-stage luminal patients (discovery cohort).**

Discovery Young Luminal | DDFS

| Model | HR | CI_lo | CI_hi | p | sig | p_fmt |
|---|---|---|---|---|---|---|
| Univariable | 2.161331 | 1.649526 | 2.831934 | 0.00000002272021 | Significant | 2.27e-08 |
| Density-adjusted | 2.219873 | 1.645527 | 2.994686 | 0.00000017840408 | Significant | 1.78e-07 |

Discovery Young Luminal | MSPI-focused C-index

| Model | C | CI_lo | CI_hi | AIC |
|---|---|---|---|---|
| MSPI only | 0.6580 | 0.5811 | 0.7345 | 459.4 |
| Density + MSPI | 0.7003 | 0.6390 | 0.7911 | 458.4 |

Discovery Young Luminal | Clinical MV Forest plot

| Variable | HR | CI_lo | CI_hi | p | p_fmt |
|---|---|---|---|---|---|
| MSPI (per SD) | 2.309 | 1.725 | 3.091 | 0.00000001828358 | 1.83e-08 |
| Stage: Stage IIa | 0.935 | 0.422 | 2.073 | 0.86862241303457 | 8.69e-01 |
| Stage: Stage IIb | 2.652 | 1.098 | 6.407 | 0.03018136985277 | 3.02e-02 |
| Systemic treatment: Chemotherapy and Endocrine treatment | 0.947 | 0.342 | 2.628 | 0.91723330172438 | 9.17e-01 |
| Systemic treatment: Endocrine treatment | 0.535 | 0.154 | 1.856 | 0.32449420178463 | 3.24e-01 |
| Systemic treatment: No systemic treatment | 0.738 | 0.232 | 2.352 | 0.60734226443632 | 6.07e-01 |
| Diagnosis period: 1998-2002 | 1.290 | 0.578 | 2.876 | 0.53405088844345 | 5.34e-01 |
| Diagnosis period: 2003-2005 | 0.852 | 0.349 | 2.078 | 0.72432879479560 | 7.24e-01 |

**Supplementary Table S9. Distribution of clinicopathological features by qMSPI quadrant in the validation cohort.**

| Characteristic | Q1 Best<br>N = 69[1] | Q2 Mixed<br>N = 84[1] | Q3 Interm<br>N = 83[1] | Q4 Worst<br>N = 69[1] | p-value[2] |
|---|---|---|---|---|---|
| **ER status** | | | | | 0.3 |
| ER+ | 51 (73.9%) | 68 (81.0%) | 69 (83.1%) | 59 (85.5%) | |
| ER- | 18 (26.1%) | 16 (19.0%) | 14 (16.9%) | 10 (14.5%) | |
| **Age** | | | | | **0.001** |
| >50 | 44 (63.8%) | 62 (73.8%) | 75 (90.4%) | 51 (73.9%) | |
| <=50 | 25 (36.2%) | 22 (26.2%) | 8 (9.6%) | 18 (26.1%) | |
| **Histological grade** | | | | | 0.059 |
| 1-2 | 59 (85.5%) | 74 (88.1%) | 81 (97.6%) | 62 (89.9%) | |
| 3 | 10 (14.5%) | 10 (11.9%) | 2 (2.4%) | 7 (10.1%) | |
| **Nodal status** | | | | | 0.3 |
| N0 | 55 (79.7%) | 69 (82.1%) | 74 (89.2%) | 55 (79.7%) | |
| N+ | 14 (20.3%) | 15 (17.9%) | 9 (10.8%) | 14 (20.3%) | |

[1]n (%)

[2]Pearson's Chi-squared test

**Supplementary Table S10. MSPI hazard ratios by ER status in the validation cohort.**

| Subgroup | N | Events | UV HR (95% CI) | UV p | MV HR (95% CI) | MV p |
|---|---|---|---|---|---|---|
| All invasive | 305 | 132 | 1.17 (0.99-1.37) | 0.061 | 1.14 (0.97-1.33) | 0.117 |
| ER-positive | 247 | 105 | 1.30 (1.09-1.55) | 0.004 | 1.27 (1.06-1.51) | 0.008 |
| ER-negative | 58 | 27 | 0.74 (0.45-1.21) | 0.235 | 0.71 (0.44-1.13) | 0.146 |

**Supplementary Table S11. Age-stratified MSPI prognostic performance in ER-positive patients from the validation cohort.**

| AgeGroup | Model | HR | CI_lo | CI_hi | p | sig |
|---|---|---|---|---|---|---|
| Age <=50 | Univariable | 2.0435781 | 1.2535738 | 3.331444 | 0.0041524927 | Significant |
| Age <=50 | Density-adjusted | 1.9940887 | 1.1742795 | 3.386238 | 0.0106310670 | Significant |
| Age 51-64 | Univariable | 1.7686488 | 1.3241677 | 2.362328 | 0.0001127539 | Significant |
| Age 51-64 | Density-adjusted | 1.7364679 | 1.2969219 | 2.324982 | 0.0002106117 | Significant |
| Age >=65 | Univariable | 0.9600516 | 0.7526956 | 1.224531 | 0.7426216565 | NS |
| Age >=65 | Density-adjusted | 0.9454690 | 0.7435363 | 1.202244 | 0.6473634464 | NS |

Validation ER+ Age-Stratified C-index

| AgeGroup | Model | C | CI_lo | CI_hi | AIC |
|---|---|---|---|---|---|
| Age <=50 (n=55) | MSPI only | 0.7185 | 0.5422 | 0.8603 | 81.3 |
| Age <=50 (n=55) | Density + MSPI | 0.7542 | 0.6372 | 0.8898 | 86.3 |
| Age 51-64 (n=92) | MSPI only | 0.6381 | 0.5301 | 0.7695 | 196.3 |
| Age 51-64 (n=92) | Density + MSPI | 0.6781 | 0.6226 | 0.8117 | 199.9 |
| Age >=65 (n=100) | MSPI only | 0.4787 | 0.4406 | 0.5913 | 510.1 |
| Age >=65 (n=100) | Density + MSPI | 0.5585 | 0.4969 | 0.6729 | 510.4 |

**Supplementary Table S12. Pooled individual patient data meta-analysis of MSPI in young patients with early luminal breast cancer.**

| Label | HR | CI_lo | CI_hi | p | n | events |
|---|---|---|---|---|---|---|
| Discovery: Young_Luminal_early | 2.161331 | 1.649526 | 2.831934 | 0.0000000227202107 | 152 | 51 |
| Validation: ER_pos_Young | 2.043578 | 1.253574 | 3.331444 | 0.0041524927151575 | 55 | 13 |
| Pooled (stratified Cox) | 2.132772 | 1.683589 | 2.701798 | 0.0000000003447686 | 207 | 64 |
